# Polygenic risk scores for asthma and allergic disease associate with COVID-19 severity in 9/11 responders

**DOI:** 10.1101/2022.08.30.22279383

**Authors:** Monika A. Waszczuk, Olga Morozova, Elizabeth Lhuillier, Anna R. Docherty, Andrey A. Shabalin, Xiaohua Yang, Melissa A. Carr, Sean A. P. Clouston, Roman Kotov, Benjamin J. Luft

## Abstract

**Background:** Genetic factors contribute to individual differences in the severity of coronavirus disease 2019 (COVID-19). A portion of genetic predisposition can be captured using polygenic risk scores (PRS). Relatively little is known about the associations between PRS and COVID-19 severity or post-acute COVID-19 in community-dwelling individuals.

**Methods:** Participants in this study were 983 World Trade Center responders infected for the first time with SARS-CoV-2 (mean age at infection=56.06; 93.4% male; 82.7% European ancestry). Seventy-five (7.6%) responders were in the severe COVID-19 category; 306 (31.1%) reported at least one post-acute COVID-19 symptom at 4-week follow-up. Analyses were adjusted for population stratification and demographic covariates.

**Findings:** The asthma PRS was associated with severe COVID-19 category (odds ratio [*OR*]=1.61, 95% confidence interval: 1.17-2.21) and more severe COVID-19 symptomatology (β=.09, *p*=.01), independently of respiratory disease diagnosis. Severe COVID-19 category was also associated with the allergic disease PRS (*OR*=1.97, [1.26-3.07]) and the PRS for COVID-19 hospitalization (*OR*=1.35, [1.01-1.82]). PRS for coronary artery disease and type II diabetes were not associated with COVID-19 severity.

**Conclusion:** Recently developed polygenic biomarkers for asthma, allergic disease, and COVID-19 hospitalization capture some of the individual differences in severity and clinical course of COVID-19 illness in a community population.

## Introduction

Coronavirus disease 2019 (COVID-19) is caused by an infection by the severe acute respiratory syndrome coronavirus (SARS-CoV-2) and remains a pandemic in the US and globally [1]. COVID-19 invades the respiratory and other crucial systems in the body, varying widely in severity, from asymptomatic presentation to severe outcomes including hospitalization or death. Moreover, at least one-third of individuals with COVID-19 develop post-acute COVID-19 syndrome [2], which consists of residual respiratory, fatigue, central nervous system, and musculoskeletal symptoms that persist at least four weeks after initial symptom onset [3, 4]. Despite vaccination efforts, the potentially severe and chronic outcomes of COVID-19 necessitate a better characterization of individuals at elevated risk for clinically significant disease outcomes. Across a range of studies, COVID-19 severity was found to be associated with clinical risk factors such as older age, male sex, smoking, respiratory illness, cardiovascular disease, and immunocompromised status [5-7]. Moreover, a person’s own genetic factors critically affect individual differences in COVID-19 disease severity [8]. Large-scale, population-based genome-wide association studies (GWAS) indicate that multiple common genetic variants can contribute to COVID-19 symptom severity after infection [9]. Genes that emerged from GWAS most strongly associated with severe COVID-19 progression are involved in immune response and lung disease pathology. The individual variants associated with disease severity in the discovery GWAS have minor effects, but cumulatively they explain a substantial proportion of genetic variation in response to the virus. Common variants can be collapsed into a polygenic risk score (PRS) to improve the prediction of at-risk patients in an independent cohort. For each individual, the number of risk alleles carried at each variant (0, 1, or 2) can be summed and weighted by its effect size, resulting in a single score of each individual’s genetic vulnerability to COVID-19 severity.

COVID-19 PRS have shown to be associated with increased risk of morbidity and mortality due to COVID-19 in several independent samples and across ancestries, with PRS outperforming individual genetic variants and providing information over and above established risk factors [10-13]. While the COVID-19 PRS was designed to capture all genes associated with the severity of COVID-19, the studies relied partially on the UK Biobank cohort, which has limited phenotypic information on COVID-19 disease and exposure status of controls, largely based on electronic medical records of laboratory test results and ICD-10 codes for COVID-19 and respiratory support. It is therefore plausible that other PRS capture genetic vulnerability to other conditions, such as asthma or coronary artery disease, that might also associate with COVID-19 symptom severity or COVID-19 clinical course, although empirical evidence is limited. For example, only one published study has reported no association between asthma PRS and severe COVID-19 [14], but no other studies to our knowledge have examined post-acute COVID-19 symptoms.

Therefore, the present study translated genetic discoveries in COVID-19 host genetics to an independent, well-characterized patient population with a full range of COVID-19 severity. Responders to the World Trade Center (WTC) attacks on September 11, 2001 are a cohort with detailed prospective health data, including information on COVID-19 infection, symptom severity, and follow-up outcomes. Our team has previously demonstrated that younger and employed responders were at a higher risk of being infected with the SARS-CoV-2 virus [15]. Among those who experienced infection, a pre-existing obstructive airway disease diagnosis predicted an elevated risk for a more severe COVID-19 presentation [2]. The risk of developing post-acute COVID-19 sequelae was similarly elevated in responders with respiratory problems as well as heart disease, over and above the risk conferred by the COVID-19 severity itself. While respiratory and other chronic conditions are associated with toxic exposures during the 9/11 disaster, some of the individual differences in post-exposure outcomes in 9/11 responders can be attributed to genetic vulnerabilities [16, 17].

Relatively little work describing genetic risk to COVID-19 severity has been conducted in community individuals with a full range of symptom severity. The associations between PRS and a common long-term outcome of post-acute COVID-19 also remain unknown. Thus, building on our previous work in the large sample of WTC responders who were infected with SARS-CoV-2, we investigated whether polygenic vulnerabilities to COVID-19 hospitalization, as well as to asthma, related allergic diseases, coronary artery disease, and type II diabetes, were associated with COVID-19 severity and post-acute COVID-19 symptoms, after adjusting for established risk factors. We investigated genetic associations with a full range of COVID-19 symptoms, as well as genetic risk for the severe COVID-19, which has the highest clinical relevance as it includes hospitalized cases. Based on previous findings, we hypothesized that PRS for COVID-19 hospitalization would be significantly associated with COVID-19 severity across a range of symptoms, and would help identify the most severe COVID-19 group. We tentatively hypothesized that PRS for asthma, allergic disease, coronary artery disease, and type II diabetes would be associated with worse COVID-19 severity outcomes and that the polygenic associations would extend to development of post-acute COVID-19 sequelae. We

## Methods

### Participants and Procedure

Participants were 983 WTC responders enrolled in a prospective cohort called the WTC Health Program which was established in 2002 and independently monitors the health of more than 13,000 WTC first responders living on Long Island, NY, USA [18]. The analytic sample comprised of all genotyped participants from an established cohort of responders with the first instance of SARS-CoV-2 infection before January 2022, with the median infection date of December 2020 [2]. The mean age of the analytic sample at infection was 56.06, SD=7.37, range= 39.79-89.04, 93.4% of the sample was male, and 82.3% had European ancestry. The analytic sample did not differ significantly from the total cohort of responders with SARS-CoV-2 infection (N=1,280) on demographic and symptom variables (all p-values for independent sample t-tests and chi-square tests >0.05).

The analytic sample included responders with a verified history of a positive polymerase chain reaction, antigenic, or antibody test for SARS-CoV-2 or positivity for antibodies to SARS-CoV-2 nucleocapsid protein according to medical or laboratory records *(N=*587), as well as responders who reported a positive laboratory test for SARS-CoV-2, but for whom documented proof of the test results was not available (*N*=396). The unverified cases did not differ significantly from verified cases on all study variables, except for lower COVID-19 severity (*t(df)*=2.30 (847.60), *p*=.02, *d*=.15). Verifiable hospitalization records at local hospitals increased the likelihood of objectively verifying the most severe responders. Analyses adjusted for verification status. All cases represent a first instance of SARS-CoV-2 infection.

Clinical information on initial and residual COVID-19 symptoms was collected irrespective of verification status through in-person questionnaires, surveys sent via email and text, electronic medical records obtained with the release of health information forms, and follow-up calls. Symptoms of COVID-19 in this study included respiratory symptoms (i.e., shortness of breath, chest pain, sore throat, congestion/runny nose, and wheezing), systemic symptoms (i.e., fever, fatigue, headache, chills, and weight loss), gastrointestinal symptoms (i.e., nausea, vomiting, and diarrhea), musculoskeletal symptoms (i.e., joint and muscle pain), central nervous system symptoms (i.e., dizziness, vertigo, loss of smell/taste, and brain fog), and psychiatric symptoms (i.e., anxiety, depression, and post-traumatic stress disorder [PTSD]).

Blood samples for genotyping were drawn at the WTC Health Program clinic as a routine part of the monitoring examination from January 2012 to January 2019. The study was approved by the Institutional Reviewer Board of Stony Brook University, and all participants provided written informed consent. For more information on the study design, see Lhuillier, Yang [2].

## Measures

### COVID-19 severity

Participants in the analytic sample were categorized into four severity groups according to their symptoms: asymptomatic (*N*=92, 9.4%), mild (*N*=378, 38.5%), moderate (*N*=408, 41.5%), and severe (*N*=75, 7.6%). This categorization was based on the NIH COVID-19 clinical spectrum updated October 2021 (National Institutes of Health, 2021). The asymptomatic category consisted of responders who reported a positive SARS-CoV-2 virologic test result without any symptoms associated with COVID-19. The mild category included responders with at least one symptom associated with COVID-19 but no shortness of breath or difficulty breathing. Moderate cases were in responders who reported shortness of breath and/or diagnosis of lower respiratory disease (pneumonia/bronchitis) during clinical assessment or imaging. These responders maintained oxygen saturation (SpO_2_) ≥ 94% on room air at sea level. Mild and moderate cases were medically managed primarily at home, even if they initially visited a healthcare facility for medical treatment and/or testing. Severe cases included responders with SpO_2_<93% on room air, respiratory rate >30 breaths/min, heart rate greater than 100 beats per minute, acute respiratory distress syndrome, septic shock, cardiac dysfunction, or an exaggerated inflammatory response in addition to pulmonary disease, or severe illness causing cardiac, hepatic, renal, central nervous system, or thrombotic disease during COVID-19. Responders were also categorized as severe if they were admitted to the hospital, or received intensive care or mechanical ventilation, or if they eventually died from COVID-19.

Two complimentary analytic variables were created: an ordinal COVID-19 severity variable on 1-4 scale corresponding to asymptomatic, mild, moderate, and severe symptoms, and a binary COVID-19 severe category variable, with asymptomatic, mild, and moderate patients in one category, and severe patients in the second category.

### Post-acute COVID-19 symptoms

Residual symptoms were defined as any COVID-19-related symptoms that lasted at least 4 weeks after symptom onset (Centers for Disease Control and Prevention, 2021b). Residual symptoms included respiratory problems (e.g., dyspnea, chest discomfort, cough, and fatigue), CNS symptoms (e.g., loss or reduction of smell/taste, mental fog, dizziness, and vertigo), and musculoskeletal complaints. In the analytic sample, N=306 (31.1%) participants reported at least one residual COVID-19 symptom. A binary analytic variable was created to compare participants with and without at least one residual symptoms.

### Covariates

Electronic medical records updated on January 1, 2020, were used to obtain date of birth to calculate age at infection, obstructive airway disease and upper respiratory disease diagnoses, lifetime posttraumatic stress disorder (PTSD) and major depressive disorder (MDD) diagnoses, and the most recent available measure of body mass index (BMI). Obstructive airway disease category includes diagnoses of asthma and bronchitis, upper respiratory disease includes chronic rhinitis and chronic sinusitis [18]. Respiratory diagnoses were determined based on systemic examination by a physician or nurse practitioner and included repeated pulmonary function tests, physical examination, and medical history. BMI scores were analyzed as continuous variable to maximize statistical power [19, 20]. Genetic data were used to obtain sex and ancestry information.

### Genetic data processing and polygenic risk scores (PRS)

Genotyping of blood samples was performed at the Genomics Shared Resource at Roswell Park Cancer Institute, using the Infinium Global Screening Array (Illumina, San Diego, CA, USA), according to manufacturer protocols. Genotypes were imputed on the Michigan Imputation Server pipeline v1.2.4, using the Haplotype Reference Consortium reference panel [21]. Before imputation, the genotypes were filtered for ambiguous strand orientation, missingness rate>5% (by marker exclusion, then by individual), Hardy-Weinberg equilibrium violation (*p*<10^−6^), and sex mismatch. After imputation, the single-nucleotide polymorphisms (SNPs) were excluded for imputation R^2^<0.5 and the average call rate below 90%. Genotype imputation was performed on 552,230 SNPs, resulting in 25,514,638 SNPs after quality control, which were used for matching to discovery GWAS variants for the final polygenic risk scoring.

PRS were created using PRSice 2.0 [22]. PRS were created by aggregating genetic variants that emerged from the GWAS of the phenotype of interest, i.e. the GWAS discovery sample. Our primary analysis used a complete list of SNPs after clumping, and their corresponding weights from GWAS discovery samples (P-value threshold=1) to incorporate more of the genome [23]. All analyses adjusted for the first ten genetic ancestry principal components (PCs). Primary analyses were conducted on participants of European ancestry due to the discovery GWAS being European; however, sensitivity analyses in the entire sample are also reported in the Supplementary Materials.

PRS for asthma and allergic disease were based on two GWAS conducted on participants of European ancestry from the UK Biobank [24]. The asthma GWAS was conducted in 90,853 individuals, 14,085 physician-diagnosed asthma cases, and 76,768 controls without asthma or allergic diseases. Asthma is heterogeneous phenotype and cases with allergic, non-allergic, mixed, and other forms of asthma were included in GWAS. The allergic disease GWAS was conducted in 102,453 individuals, 25,685 with doctor-diagnosed hay fever, allergic rhinitis, eczema, or allergic asthma cases, and 76,768 controls without asthma or allergic disease.

The coronary artery disease PRS was based on a GWAS meta-analysis conducted in 122,733 coronary artery disease cases and 424,528 controls from the UK Biobank and the CARDIoGRAMplusC4D Consortium [25]. Cases included patients with coronary artery disease diagnosis or treatment indicated in their electronic medical record or self-reported heart attack/myocardial infarction, coronary angioplasty, or heart bypass. The control group excluded participants who reported that their mother, father, or sibling suffered from heart disease. The type II diabetes PRS was based on a GWAS meta-analysis in 62,892 type II diabetes cases and 596,424 controls of European ancestry [26].

PRS for COVID-19 severity were based on summary statistics from two GWAS meta-analyses conducted by the COVID-19 Host Genetics Initiative [9] release 6, dated June 15, 2021. This initiative is an international collaboration spanning 61 studies from 24 countries. The first GWAS was performed in 2,085,803 individuals, contrasting 24,274 patients hospitalized due to COVID-19 with 2,061,529 population controls presumed to be COVID-19 negative (COVID-PRS hospitalized vs. population). The second GWAS was conducted in 87,671 patients with COVID-19, contrasting 14,480 hospitalized patients with 73,191 non-hospitalized patients (COVID-PRS hospitalized vs. not-hospitalized).

### Analytic approach

Linear regression was used to test whether PRS is associated with ordinal COVID-19 severity. Logistic regression was used to test whether PRS is associated with the severe COVID-19 category classification (severe vs. moderate/mild/asymptomatic), and the presence of post-acute COVID-19 symptoms. Continuous variables were standardized prior to analysis by transforming them to Z scores with a mean of zero and standard deviation of one. Models were adjusted for the first ten principal components of the population structure, COVID-19 verification status, age, and sex. Models testing asthma and allergic disease PRS were further adjusted for obstructive airway disease and upper respiratory disease diagnoses to ensure that associations do not merely reflect respiratory illness status. A supplementary analysis further adjusting for PTSD and MDD diagnoses was conducted for asthma PRS, to test the potential role of mental health in the associations. Likewise, models testing PRS for coronary artery disease and type II diabetes further adjusted for BMI. Models testing associations with residual symptoms additionally adjusted for COVID-19 severity. Separate models were conducted for each PRS. The primary analyses were conducted in participants of European ancestry. Given that COVID-19 GWAS were completed using multi-ancestry participants, for sensitivity testing and inclusivity results are also reported for participants of all ancestries in Supplementary Materials. All analyses used nominal p-value threshold <.05 and were conducted in SPSS version 28 and R version 4.1.0.

## Results

Descriptive statistics for the European ancestry sample are reported in Table 1. Many responders (*N=*57, 7.0%) had severe COVID-19. In total, 247 (30.4%) responders reported at least one residual COVID-19 symptom. Correlations between PRS are reported in Supplementary Table S1.

**Table 1.** Descriptive statistics for participants of European ancestry.

|  |  |
| --- | --- |
| N total | 813 |
| N male (%) | 772 (95.0%) |
| Mean age at infection (SD), range | 56.06 (7.37), 39.79-89.04 |
| N OAD diagnosis (%) | 312 (38.4%) |
| N URD diagnosis (%) | 525 (64.6%) |
| N PTSD and/or MDD diagnosis (%) | 124 (15.3%) |
| Mean BMI (SD), range | 31.34 (5.08), 19.48-62.18 |
| N COVID-19 severity (%) |  |
| Asymptomatic | 78 (9.6%) |
| Mild | 320 (39.4%) |
| Moderate | 332 (40.8%) |
| Severe | 57 (7.0%) |
| Missing | 26 (3.2%) |
| N any residual symptoms | 247 (30.4%) |

*Notes:*
OAD – Obstructive airway disease; URD – upper respiratory disease; PTSD-Posttraumatic stress disorder, MDD – Major depressive disorder, BMI – body mass index; COVID-19 – coronavirus disease 2019; SD - standard deviation.

The PRS for asthma was significantly associated with COVID-19 severity in the European ancestry subsample (β=.09, *p*=.01), adjusting for co-occurring obstructive airway disease diagnoses and upper respiratory disease diagnoses, Table 2. Likewise, the PRS for asthma was associated with COVID-19 severe category (*OR*=1.61, [1.17-2.21]). Allergic disease PRS showed a similar pattern, significantly associating the COVID-19 severe category (*OR*=1.97, [1.26-3.07]). Associations with residual COVID-19 symptoms were not significant. There were no significant associations between coronary artery disease or type II diabetes PRS, and COVID-19 phenotypes (Table 3). The COVID-19 PRS based on the GWAS contrasting hospitalized cases vs. controls was significantly associated with the COVID-19 severe category in sample with European ancestry (*OR*=1.35, [1.01-1.82]), Table 4. Conversely, the COVID-19 PRS based on the GWAS contrasting hospitalized cases vs. non-hospitalized cases did not reach significance in the European ancestry subsample. Moreover, none of the COVID-19 PRS were associated with the full range of COVID-19 severity, nor the presence of any residual symptoms.

**Table 2.** Associations between allergic disease and asthma PRS and COVID-19 severity and residual symptoms in participants of European ancestry.

|  | COVID-19 severity | COVID-19 severe category | Any residual symptoms |
| --- | --- | --- | --- |
| PRS: Asthma | $\beta=.09, p=.01$ | **OR=1.61**<br>(CI: 1.17-2.21), $p<.01$ | OR=1.07<br>(CI: .90-1.27), $p=.43$ |
| PRS: Allergic disease | $\beta=.09, p=.06$ | **OR=1.97**<br>(CI: 1.26-3.07), $p<.01$ | OR=.89<br>(CI: .70-1.13), $p=.34$ |

*Notes:*
OR: Odds ratio; CI: 95% confidence interval; PRS: polygenic risk score; COVID-19: coronavirus disease 2019. All models are adjusted for the first ten principal components of the population structure, verification status, age at infection, sex, obstructive airway disease diagnosis, upper respiratory disease diagnosis. Models with residual symptoms as a dependent variable were additionally adjusted for COVID-19 severity. Associations between asthma PRS and COVID-19 outcomes were not affected by lifetime PTSD and MDD diagnoses, see Supplementary Table S6.

**Table 3.**
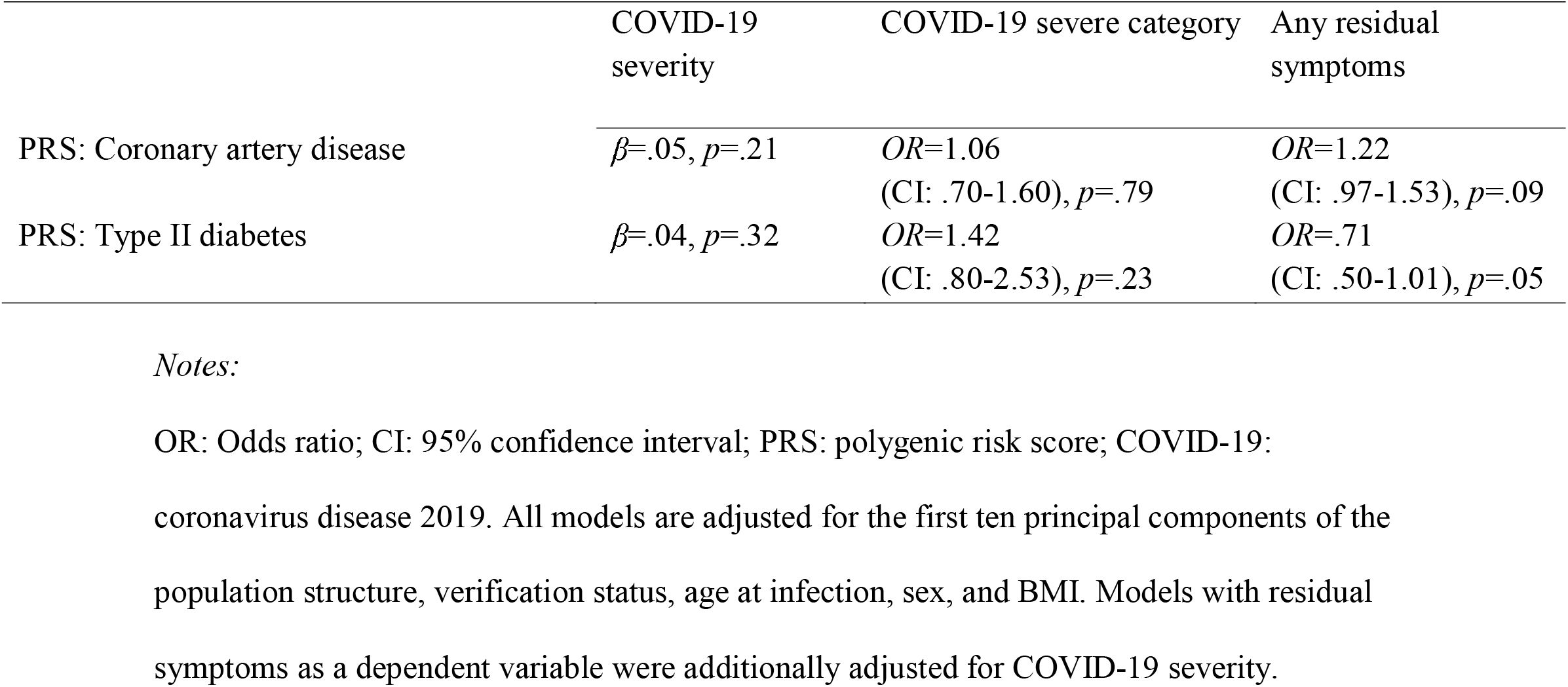
Associations between coronary artery disease and type II diabetes PRS and COVID-19 severity and residual symptoms in participants of European ancestry.

**Table 4.** Associations between COVID-19 PRS and COVID-19 severity and residual symptoms in participants of European ancestry.

|  | COVID-19 severity | COVID-19 severe category | Any residual symptoms |
| --- | --- | --- | --- |
| PRS: COVID-19 hospitalized vs. controls | $\beta=.03, p=.40$ | **OR=1.35**<br>(CI: 1.01-1.82), $p=.04$ | OR=1.11<br>(CI: .93-1.33), $p=.24$ |
| PRS: COVID-19 hospitalized vs. not-hospitalized | $\beta=.05, p=.23$ | OR=1.27<br>(CI: .95-1.69), $p=.12$ | OR=1.08<br>(CI: .90-1.29), $p=.41$ |

*Notes:*
OR: Odds ratio; CI: 95% confidence interval; PRS: polygenic risk score; COVID-19: coronavirus disease 2019. All models are adjusted for the first ten principal components of the population structure, verification status, age at infection, and sex. Models with residual symptoms as a dependent variable were additionally adjusted for COVID-19 severity.

### Sensitivity analyses

Descriptive statistics for total sample with participants of all ancestries are reported in Supplementary Table S2. In the total sample, adjusting for respiratory disease diagnoses, the PRS for asthma was significantly associated with COVID-19 severity (β=.08, *p*=.01) and COVID-19 severe category (*OR*=1.53, [1.15-2.03]), and the allergic disease PRS was associated with the COVID-19 severe category (*OR*=1.86, [1.25-2.77]), Supplementary Table S3. There were no significant associations between coronary artery disease or type II diabetes PRS, and COVID-19 phenotypes, Supplementary Table S4. The COVID-19 PRS based on the GWAS contrasting hospitalized cases vs. controls was significantly associated with the COVID-19 severe category in the whole sample (*OR*=1.42, [1.11-1.82]), Supplementary Table S5. Likewise, the COVID-19 PRS based on the GWAS contrasting hospitalized cases vs. non-hospitalized cases was also significantly associated with COVID-19 severe category in the total sample (*OR*=1.33, [1.04-1.69]). Accounting for PTSD and MDD diagnoses did not alter associations between asthma PRS and COVID-19 outcomes, see Supplementary Table S6.

## Discussion

The current study reports associations between recently developed polygenic biomarkers and COVID-19 severity in a community-based population of responders to the WTC disaster who had COVID-19. We found that the PRS for asthma, allergic diseases, and COVID-19 severity were elevated in the severe COVID-19 category responders.. Asthma PRS was additionally associated with COVID-19 symptomatology across the full spectrum of COVID-19 severity. No significant associations were observed for PRS for coronary artery disease and type II diabetes. Moreover, PRS were not associated with post-acute COVID-19 sequelae. These findings add to the growing body of evidence to suggest that existing polygenetic profiles could be informative about the risk for severe COVID-19 presentation, despite the small effect sizes.

The current study is the first to report significant associations between asthma PRS and allergic disease PRS, and COVID-19 severity. A previous study in the UK Biobank found no association between asthma PRS and severe COVID-19 [14]. However, the direction of association was positive, and asthma diagnosis was significantly associated with COVID-19 severity (*OR*=1.39), in line with epidemiological and clinical evidence [7]. In contrast to Zhu, Hasegawa [14], in our subsample with European ancestry, the PRS for asthma was associated with a 1.61-fold increase in the odds of classification into the most severe COVID-19 category, as well as with the full range of COVID-19 symptom severity, independently of respiratory diagnoses. Our focus on patients with a wide range of COVID-19 symptoms, in contrast with Zhu, Hasegawa [14] focus on hospitalized cases with severe COVID-19, might be a strength of the current study that explains different findings. Other potential explanations include demographic differences between the samples, data collection at different phases of the pandemic, and differences in sample sizes.

Allergic disease PRS based on the related GWAS of cases with doctor-diagnosed hay fever, allergic rhinitis, eczema, or allergic asthma doubled the odds of classification into the most severe COVID-19 category in participants of European ancestry (*OR*=1.97), adjusting for demographic covariates and respiratory diagnoses. Taken together, these results are consistent with the evidence for the role of immune function genes implicated in COVID-19 severity [8]. It is possible that asthma and allergic sensitization contribute to the pathobiology of severe COVID-19, such as expression of the SARS-CoV-2 receptor ACE2 [27, 28], and that asthma and allergic disease PRS capture these effects. Another possibility is that both asthma and atopic disease have a proclivity toward a TH2 immune response and thus may contribute to a TH1 or TH2 imbalance and a poorer outcome [29]. Finally, despite the substantial genetic correlation between asthma and mental health [30], responders’ history of PSTD and MDD did not explain the association between asthma PRS and COVID-19 severity.

Contrary to our hypotheses, PRS for coronary artery disease and type II diabetes were not associated with any COVID-19 outcomes. Cardiovascular and other obesity-related disorders have previously been highlighted as risk factors for severe COVID-19 presentation and residual symptoms [7, 31], including in our sample [2]. However, our findings suggest that genetic vulnerability to these conditions might not contribute prominently to COVID-19 severity. A study found a significant association between BMI PRS and higher risk of severe COVID-19 outcomes such as hospitalization; however, the effect sizes were small (*OR*=1.14) [32] and were below effect sizes the present study was powered to detect. Furthermore, the previous study did not find significant relationships between COVID-19 outcomes and PRS for BMI-adjusted waist-to-hip ratio and BMI-adjusted waist circumference, indicating an overall weak association between genetic vulnerability to high weight and COVID-19 severity, consistent with the current findings for PRS developed for coronary artery disease and type II diabetes.

The PRS based on the largest GWAS to date comparing hospitalized COVID-19 cases to healthy controls [9] was significantly associated with a 1.35-fold increase in the odds of having had severe COVID-19. This finding agrees with previous studies based on large cohorts such as the UK Biobank which found that COVID-19 PRS were associated with severe COVID-19 outcomes [10-12]. For example, Nakanishi, Pigazzini [10] reported that COVID-19 PRS predicted death or severe respiratory failure (OR=1.70). Similarly, Horowitz, Kosmicki [11] reported that a PRS based on the top six SNPs associated with COVID-19 predicted a 1.38-fold increased risk of hospitalization and a 1.58-fold increased risk of severe disease in European ancestry participants, with results replicated in other ancestries. Importantly, COVID-19 severity PRS were not associated with the full spectrum of COVID-19 symptoms in our population and did not associate with residual post-acute COVID-19 syndrome. This finding adds novel evidence that the COVID-19 PRS specifically captures genetic vulnerability to a severe response to SARS-CoV-2 infection.

### Strengths and Limitations

The strengths of the current study include a large sample with well-characterized COVID-19 symptomatology, follow-up information on post-acute COVID-19 sequelae, and application of PRS based on the largest discovery samples available for each disease. Nonetheless, we also note several limitations. First, although we demonstrated that genotyped patients did not differ significantly from non-genotyped patients, the specific inclusion and exclusion criteria might result in selection bias, contributing to the broader non-representativeness of the WTC cohort to the general population. The proportion of individuals with ancestry admixture in the WTC cohort is too small to allow genetic analyses in non-Northern European ancestries. Nonetheless, for transparency, we opted to report analyses in the total sample as well as the European ancestry subsample, and results are notably similar. In either sample, we were underpowered to detect very small associations. Relatedly, the WTC responder population is primarily male, and the number of females was insufficient to explore sex differences. Moreover, we were underpowered to stratify the sample by age, respiratory illness, or other characteristics related to COVID-19 severity. We have adjusted for these covariates in our analyses, but acknowledge that the findings arising from the current study may not generalize to other populations.

Second, some participants were included based on infection information that was not independently verified by the study team. However, these participants reported testing positive for SARS-CoV-2 through one of the available testing methods and did not differ from verified participants on any characteristics except COVID-19 severity, which is due to verified records being available for hospitalized responders, suggesting that misclassification was likely to be minimal. The COVID-19 verification status was included as a covariate in all analyses. Third, different viral strains emerged and vaccination status changed over the course of this study. These factors can affect infection susceptibility and COVID-19 disease severity. For example, COVID-19 vaccines are very effective in lowering infection and disease severity [33, 34], which in turn could decrease the role of host genetics in COVID-19 outcomes, attenuating observed PRS associations. Moreover, in vaccinated individuals, PRS associations with COVID-19 severity could instead reflect vulnerability to vaccine breakthroughs. We did not have sufficient data to investigate these issues, which likely have contrasting effects on our results, and limit their comparability to studies conducted at different points in the pandemic. Future studies of PRS are needed to model complex interactions between these factors.

Fifth, the current study used asthma PRS based on a GWAS of a broadly defined asthma. However, asthma is a complex and heterogeneous condition, with allergic and non-allergic asthmas demonstrating partially different genetic etiologies [24, 35, 36]. Accordingly, PRS for specific asthma subtypes based on allergy and age-at-onset have been shown to differentially associate with outcomes such as BMI [37]. Thus, future work should replicate and clarify whether observed associations between asthma PRS and COVID-19 severity differ across asthma subtypes.

## Conclusions

The current translational study found that PRS for asthma, allergic disease, and COVID-19 hospitalization were associated with an increased risk for severe COVID-19 presentation among community-based individuals infected with SARS-CoV-2. Asthma PRS was also associated with higher COVID-19 severity across the full range of COVID-19 symptoms. The current effect sizes were small, but with additional research, known genetic vulnerability factors might help to identify individuals at an increased risk for a severe course of COVID-19 disease.

## Data Availability

Data file is available at synapse.org (https://www.synapse.org/#!Synapse:syn44590167, doi:10.7303/syn44590167) and uploaded alongside the preprint on medRxiv.

## Acknowledgements

We thank participants for their commitment to the project

## Data Availability Statement

Data file is available at synapse.org (https://www.synapse.org/#!Synapse:syn44590167, doi:10.7303/syn44590167)

## Supporting Information

**Table S1. Associations among PRS used in the study**

**Table S2. Descriptive statistics for participants of all ancestries**

**Table S3. Associations between allergic disease and asthma PRS and COVID-19 severity and residual symptoms in participants of all ancestries**

**Table S4. Associations between coronary artery disease and type II diabetes PRS and COVID-19 severity and residual symptoms in participants of all ancestries**

**Table S5. Associations between COVID-19 PRS and COVID-19 severity and residual symptoms in participants of all ancestries**

**Table S6. Mental health and associations between COVID-19 outcomes and PRS for asthma in European ancestry participants**

