## Supplementary materials for "Polygenic risk scores for asthma and allergic disease associate with COVID-19 severity in 9/11 responders"

Supplementary Table 1 –Associations among PRS used in the study

|  | PRS: Asthma | PRS: Allergic disease | PRS: Coronary artery disease | PRS: Type II diabetes | PRS: COVID-19 hospitalized vs. controls | PRS: COVID-19 hospitalized vs. not-hospitalized |
| --- | --- | --- | --- | --- | --- | --- |
| PRS: Asthma | 1 | .42* | .02 | .04 | .02 | -.01 |
| PRS: Allergic disease | .43* | 1 | -.03 | -.01 | .02 | .00 |
| PRS: Coronary artery disease | .02 | -.02 | 1 | .16* | .09* | .10* |
| PRS: Type II diabetes | .04 | -.01 | .16* | 1 | .12* | .03 |
| PRS: COVID-19 hospitalized vs. controls | .02 | .03 | .06 | .10* | 1 | .63* |
| PRS: COVID-19 hospitalized vs. not-hospitalized | .01 | .01 | .09* | .02 | .65* | 1 |

*Note:* Association in full sample reported below diagonal, associations in European ancestry reported above diagonal. Partial correlations adjusted for the first ten principal components of the population structure are reported.

* indicates *p*<.05

Supplementary Table 2 – Descriptive statistics for participants of all ancestries

| N total | 983 |
| --- | --- |
| N male (%) | 918 (93.4%) |
| Mean age at infection (SD), range | 56.03 (7.37), 39.79-89.04 |
| N OAD diagnosis (%) | 372 (37.8%) |
| N URD diagnosis (%) | 627 (63.8%) |
| N PTSD and/or MDD diagnosis (%) | 146 (14.9%) |
| Mean BMI (SD), range | 31.31 (5.09), 19.48-62.18 |
| N COVID-19 severity (%) |  |
| Asymptomatic | 92 (9.4%) |
| Mild | 378 (38.5%) |
| Moderate | 408 (41.5%) |
| Severe | 75 (7.6%) |
| Missing | 30 (3.1%) |
| N any residual symptoms | 306 (31.1%) |

*Notes:*

OAD – Obstructive airway disease; URD – upper respiratory disease; BMI – body mass index; COVID-19 – coronavirus disease 2019; SD - standard deviation.

Supplementary Table 3– Associations between allergic disease and asthma PRS and COVID-19 severity and residual symptoms in participants of all ancestries.

|  | COVID-19 severity | COVID-19 severe category | Any residual symptoms |
| --- | --- | --- | --- |
| PRS: Asthma | ***β*=.08, *p*=.01** | ***OR*=1.53 (CI:1.15-2.03), *p*<.01** | *OR*=1.11 (CI:.95-1.29), *p*=.20 |
| PRS: Allergic disease | *β*=.09, *p*=.05 | ***OR*=1.86**  **(CI: 1.25-2.77), *p*<.01** | *OR*=.94  (CI: .75-1.18), *p*=.59 |

*Notes:*

OR: Odds ratio; CI: 95% confidence interval; PRS: polygenic risk score; COVID-19: coronavirus disease 2019. All models are adjusted for the first ten principal components of the population structure, verification status, age at infection, sex, obstructive airway disease diagnosis, upper respiratory disease diagnosis. Models with residual symptoms as a dependent variable were additionally adjusted for COVID-19 severity.

Supplementary Table 4 – Associations between coronary artery disease and type II diabetes PRS and COVID-19 severity and residual symptoms in participants of all ancestries.

|  | COVID-19 severity | COVID-19 severe category | Any residual symptoms |
| --- | --- | --- | --- |
| PRS: Coronary artery disease | *β*=.05, *p*=.24 | *OR*=1.11 (CI:.77-1.61), *p*=.57 | *OR*=1.19 (CI:.96-1.47), *p*=.12 |
| PRS: Type II diabetes | *β*=.07, *p*=.27 | *OR*=1.42  (CI: .84-2.40), *p*=.19 | *OR*=.72  (CI: .53-1.00), *p*=.05 |

*Notes:*

OR: Odds ratio; CI: 95% confidence interval; PRS: polygenic risk score; COVID-19: coronavirus disease 2019. All models are adjusted for the first ten principal components of the population structure, verification status, age at infection, sex, and BMI. Models with residual symptoms as a dependent variable were additionally adjusted for COVID-19 severity.

Supplementary Table 5 – Associations between COVID-19 PRS and COVID-19 severity and residual symptoms in participants of all ancestries.

|  | COVID-19 severity | COVID-19 severe category | Any residual symptoms |
| --- | --- | --- | --- |
| PRS: COVID-19 hospitalized vs. controls | *β*=.04, *p*=.22 | ***OR*=1.42 (CI:1.11-1.82), *p*=.01** | *OR*=1.08  (CI: .92-1.26), *p*=.36 |
| PRS: COVID-19 hospitalized vs. not-hospitalized | *β*=.05, *p*=.15 | ***OR*=1.33 (CI:1.04-1.69), *p*=.02** | *OR*=1.05 (CI:.89-1.24), *p*=.55 |

*Notes:*

OR: Odds ratio; CI: 95% confidence interval; PRS: polygenic risk score; COVID-19: coronavirus disease 2019. All models are adjusted for the first ten principal components of the population structure, verification status, age at 9/11, and sex. Models with residual symptoms as a dependent variable were additionally adjusted for COVID-19 severity.

Supplementary Table 6 –Mental health and associations between COVID-19 outcomes and PRS for asthma in European ancestry participants

|  | COVID-19 severity | COVID-19 severe category | Any residual symptoms |
| --- | --- | --- | --- |
| Model 1, Asthma PRS  Mental health excluded | ***β*=.09, *p*=.02** | ***OR*=1.50**  **(CI: 1.06-2.12), *p*=.02** | *OR*=1.10  (CI: .91-1.32), *p*=.33 |
| Model 2, Asthma PRS  Mental health adjusted | ***β*=.09, *p*=.03** | ***OR*=1.60**  **(CI: 1.16-2.20), *p*<.01** | *OR*=1.13  (CI: .97-1.32), *p*=.13 |

*Notes:*

OR: Odds ratio; CI: 95% confidence interval; PRS: polygenic risk score; COVID-19: coronavirus disease 2019. All models are adjusted for the first ten principal components of the population structure, verification status, age at infection, sex, obstructive airway disease diagnosis, upper respiratory disease diagnosis. Models for residual symptoms were additionally adjusted for COVID-19 severity.

Model 1 - Subsample without PTSD and/or MDD diagnostic history (N=124 excluded).

Model 2 - Total sample, additional adjustment for PTSD and/or MDD diagnostic history.
